# Functional neurological symptoms occur commonly in healthy adults: implications for the pathophysiology of FND

**DOI:** 10.64898/2026.02.26.26347208

**Authors:** David D.G. Palmer, Mark J. Edwards, Jason B. Mattingley

## Abstract

**Objectives:** Functional neurological symptoms which do not meet clinical definitions of functional neurological disorder (FND) are common in clinical practice. Understanding the distinction between these ‘benign’ functional symptoms and FND is crucial in defining FND as an entity for study, and as a clinical syndrome. We aimed to measure the frequency of functional symptoms in people who do not have FND.

**Methods:** A survey was administered to 95 clinicians who attended an international conference on FND. Participants were asked to report the occurrence and characteristics of experiences with features of functional sensory or motor symptoms, or dissociation.

**Results:** Of the 95 people who responded to the survey, 57.4% reported having experienced any functional symptoms, and 47.9% reported having experienced functional motor or sensory symptoms. The symptoms reported were generally short-lived and caused only mild distress and disruption. Most respondents who reported having experienced a functional symptom reported having had multiple events through their lives.

**Interpretation:** The results suggest that the lifetime occurrence of functional neurological symptoms is at least two orders of magnitude higher than the prevalence of FND. The high prevalence of functional symptoms in people who have never had FND challenges the common assumption that the occurrence of functional neurological symptoms is synonymous with FND. We propose that FND is better conceived of as a failure of the mechanisms by which functional neurological symptoms resolve, rather than the occurrence of functional symptoms per se. This reconceptualization implies new research directions for the underlying aetiology of FND.

## Introduction

Functional neurological disorder (FND) is a condition characterised by the occurrence of severe and persistent functional neurological symptoms. Descriptions of FND often treat functional neurological symptoms (referred to hereafter as functional symptoms) as synonymous with FND. In clinical practice, however, neurologists frequently encounter mild functional symptoms which they do not diagnose as FND, and functional signs are often identified on examination in the absence of symptoms.^1^ Moreover, experiences which have the defining features of functional symptoms are observable in normal life and—more publicly—in sportspeople and performers, where they are referred to by names including ‘choking’, ‘the twisties’, and ‘target panic’.^2^

Functional symptoms are neurological symptoms which are generated by abnormal brain processing.^6^ They are distinctive in their striking and paradoxical variation in intensity with symptom-focused attention or effort to overcome the symptom. Typically, greater levels of symptom-focused attention are associated with increases in symptom-severity, and direction of selective attention elsewhere causes reduction or complete remission of the symptom.^7,8^ Functional symptoms encompassing almost all categories of neurological symptoms are seen, including abnormalities of movement, sensation (both somatosensation and the special senses), consciousness, and cognition.^9^

Functional symptoms are often seen as the hallmark of FND; therefore, the possibility that they might occur in many more people than are ever diagnosed with FND has the potential to change our view of the underlying dysfunction in people with FND. Given the importance of this question to understanding the aetiology of FND, we aimed to test the hypothesis that in a healthy population, the proportion who had experienced functional symptoms would be high, and that these symptoms would, on average, be mild and short-lived. Although some indirect observations have been reported which support this hypothesis,^3–5^ the frequency of functional symptoms in people who do not have FND has not yet been explicitly studied.

Retrospective classification of symptoms as functional or not is challenging, and in lay groups would likely be impossible—particularly since the symptoms we are interested in might be transient and mild. To mitigate this difficulty, we chose to study FND experts, since their knowledge of the nature of functional symptoms should give them the best chance of identifying those symptoms in themselves. We found this group by limiting inclusion to people who attended the 5^th^ International Conference on Functional Neurological Disorders. People were excluded if they had ever been diagnosed with FND. Participants completed a survey asking them to report the occurrence of experiences with the features of functional motor symptoms, functional sensory symptoms, or dissociation (formulated here as being within the same spectrum of experience as dissociative seizures) at any time. Here, we present the results of that survey with reference to our hypothesis that— assuming FND represents a failure of the mechanisms by which functional symptoms resolve—mild, self-limited functional symptoms should be common in people who do not have FND.

## Materials and methods

The survey was administered over the course of the 5^th^ International Conference on Functional Neurological Disorders held in Verona, Italy in June 2024. Inclusion was limited to attendees of the conference. Participants were recruited through posters at the conference with QR codes linked to the survey; through word of mouth, with people able to share the link to the survey with others; and by opportunistic direct conversation, with people completing the survey on an iPad.

An instrument to measure the past occurrence of functional symptoms, the Functional Neurological Experiences Questionnaire (FNEQ) was developed by the three authors, two of whom are neurologists with subspecialty expertise in FND (DP and ME). It was administered using the REDCap electronic data capture platform,^15,16^ and participants responded anonymously, providing age, gender, biological sex, and profession as demographic information. Participants were also asked to confirm that they had never been diagnosed with FND.

The FNEQ asks participants about the occurrence of motor symptoms or sensory symptoms compatible with functional symptoms at any time in life, and whether they have experienced dissociation (derealization/depersonalisation) at any time in their life. The questions are worded to be as specific for functional symptoms as possible, for example, “Have you ever had trouble moving a body part in the way you wanted to (including swallowing) which got worse the harder you tried, or seemed to work better when you were not concentrating on it?” Participants who indicate that they have experienced any of these symptoms are asked the number of times they have experienced symptoms of that type, the longest duration of symptoms of that type they have experienced, and the maximum degree of impact that the symptoms have had on their social, educational, and occupational participation. Responses to these questions are made as selections from an ordered list, and it takes most people between one and three minutes to complete. A full transcript of the FNEQ is provided as supplementary material.

The study was approved by The University of Queensland’s Health Research Ethics Committee with project number 2024/HE000730.

Data analyses were performed using Python. Descriptive statistics for the pooled participant responses are presented. Where responses are normally distributed, continuous data are presented as means with standard deviations. Non-normally distributed data are presented as medians with lower and upper quartiles.

## Data availability

Data and analysis scripts are available as a Github repository at https://github.com/ddgpalmer/Functional-neurological-symptoms-in-healthy-adults-without-FND---Data-and-Analysis.git.

## Results

In total, 95 of the 716 in-person conference attendees completed the survey. None of the respondents had been diagnosed with FND. The mean age of respondents was 40.5 years (SD 10.5, range 21 – 65). 76% of respondents were female, 21% were male, and 3% described their gender in other categories. 93% of respondents were professional clinicians, 5% were scientists or research students, and 2% other professions.

Of the respondents, 20.2% endorsed having experienced functional motor symptoms at any time, 37.9% endorsed having had functional sensory symptoms, and 30.5% endorsed having experienced dissociation. In total, 57.4% of respondents endorsed having experienced any of these symptoms, with 47.9% having experienced either a functional motor or sensory symptom.

Figure 1 presents the proportion of respondents who had ever experienced each symptom category, the duration of the longest event a participant had experienced, the number of events they had experienced, and the maximal life-impact of the symptoms. For maximum symptom duration, the median response for motor symptoms was ‘A few minutes’, with upper and lower quartiles of ‘Less than a minute’ and ‘Less than one hour’. For sensory symptoms, the median response was ‘Less than one hour,’ with quartiles of ‘A few minutes’ and ‘More than an hour, but less than a day’. For dissociation, the median was ‘A few minutes’ with quartiles of ‘Less than a minute’ and ‘More than a few minutes, but less than an hour’.

**Figure 1.**
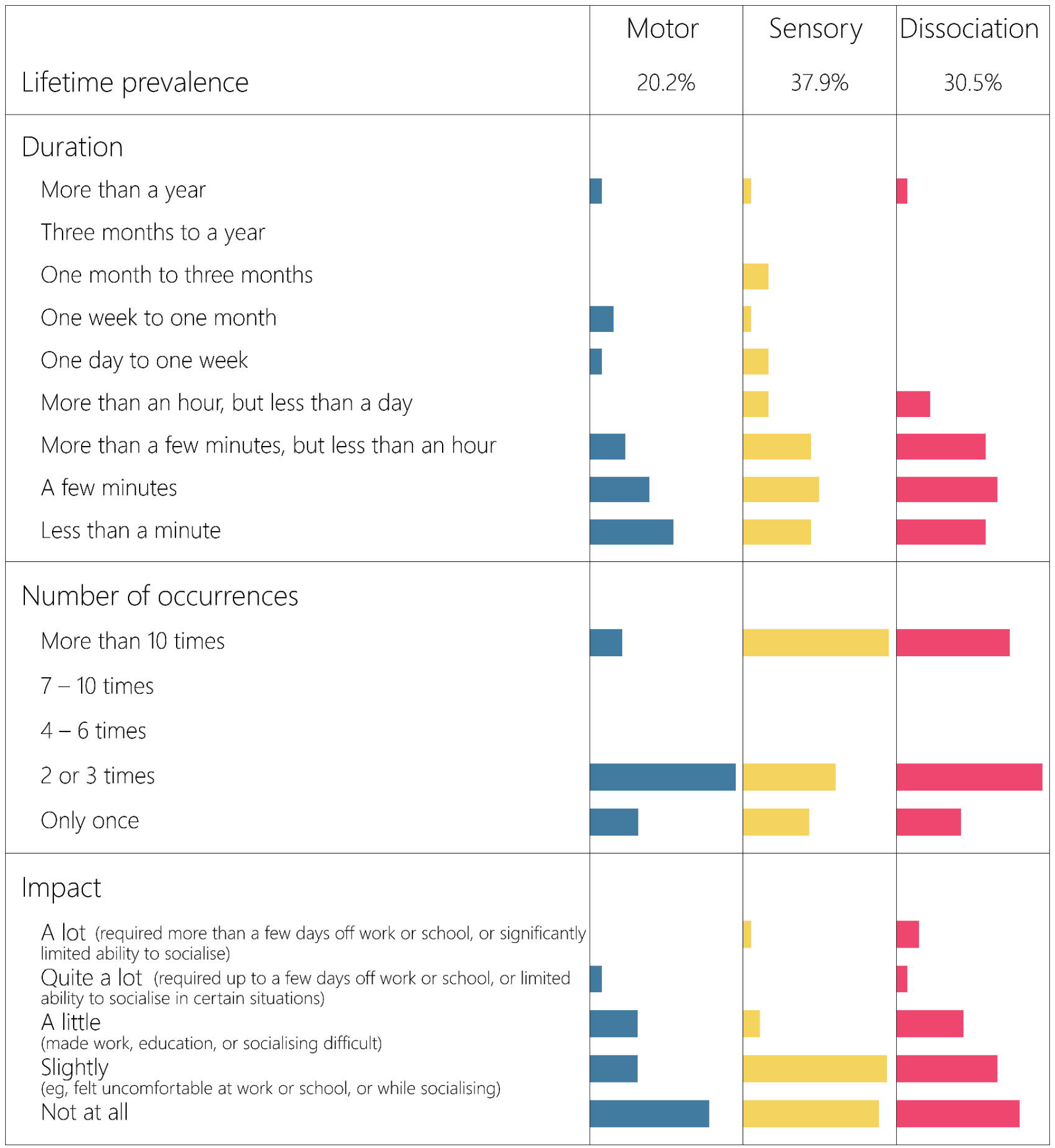
Characteristics of Functional Neurological Symptoms Experienced by Respondents. Coloured areas represent the proportion of participants reporting each category or worse. Bars represent the number of responses per category.

When reporting the number of occurrences of a symptom-type they had ever experienced, participants were instructed to consider only the occurrence of separate symptoms, and to disregard the same symptom coming and going during one episode. For motor symptoms, the median response category for distinct episodes of symptoms was ‘2 – 3,’ with lower and upper quartiles of ‘2 – 3,’ and ‘7 – 10’. For sensory symptoms, the median response was ‘4 – 6,’ with quartiles of ‘2 – 3,’ and ‘More than 10’. For dissociation, the median was ‘4 – 6,’ with quartiles of ‘2 – 3,’ and ‘7 – 10’.

For the maximal impact of symptoms on participants’ lives, the median response was ‘Not at all’ with lower and upper quartiles of ‘Not at all’ and ‘A little’. For sensory symptoms, the median response was ‘Slightly’, with lower and upper quartiles of ‘Not at all’ and ‘Slightly’. For dissociation, the median response was ‘Slightly’ with quartiles of ‘Not at all’ and ‘A little’.

## Discussion

We aimed to describe the frequency of functional symptoms in people who do not have FND. By sampling a group who are experts in the identification of functional symptoms and using an instrument worded to maximise specificity, we generated a plausible estimate of the proportion of people who do not have FND who have experienced functional symptoms. It is likely that the results represent, if anything, an underestimate of the frequency of symptoms, since mild and brief symptoms are easily forgotten.

Our results show that functional symptoms occur commonly in people who do not have FND. Over half of survey respondents indicated that they had experienced at least one functional neurological symptom in the past. The most common functional symptoms were sensory, occurring in 38% of respondents, with motor symptoms reported by 20% of respondents, and 31% of respondents reporting having experienced symptoms of dissociation (depersonalization and/or derealisation). Participants who reported past functional symptoms generally reported having had symptoms multiple times in their lives, and overall, the symptoms they reported were brief and non-distressing.

In addition to the presence of functional symptoms, current definitions of FND also require the presence of distress or loss of function due to the condition.^6,12,13^ This criterion is rooted in the tradition of psychiatric diagnosis, and optimizes the definition of FND to identify people who would benefit from treatment for their symptoms. However, it is insufficient in defining the occurrence of a distinct biological entity. Many unrelated physical and mental factors contribute to the distress and limitation of function that a person experiences from a symptom, and these are unlikely to be a defining feature of the brain dysfunction underlying FND. For research purposes, more robust distinctions need to be made between those who have the disorder and those who do not.

Based on the results of this survey, the number of people who have experienced functional symptoms outweighs the number of people who have FND by at least two orders of magnitude.^14^ This supports the need for criteria beyond the occurrence of functional symptoms in defining FND. It also supports the Bayesian brain account of functional symptoms, where mild functional symptoms should be expected to occur from time to time even in well-functioning nervous systems.^10^

Given the premises that optimally functioning brains should be expected to develop functional symptoms,^10^ and that FND is uncommon,^14^ we propose that FND might best be seen as a failure of the mechanisms by which functional symptoms resolve: not as a condition which causes the development of functional symptoms. If this is the case, we should expect that the majority of functional symptoms that occur in otherwise healthy people resolve spontaneously, with only a minority evolving into FND.

Defining FND as a failure of the mechanisms which repair functional symptoms accords with our observation that functional symptoms are markedly more common than FND, and fits the distinction tacitly made in clinical practice between ‘benign’ functional symptoms and FND. It also avoids being forced to accept the counterintuitive suggestion that the underlying difference between people who develop FND and people who do not is a mechanism which determines their emotional reaction to, or physical limitation from, functional symptoms, not a factor related to the symptoms themselves. In practice, this definition can be operationalised by defining FND as the occurrence of functional symptoms which are persistent and more than trivial in severity, which is consistent with the practice of many specialists in the area. Other features which are seen in people with FND, but not in these benign functional symptoms, such as the tendency for one symptom to spread or evolve into another, could also be seen as markers of inadequate or malfunctioning repair mechanisms for functional symptoms. Formalising these features as nosological criteria would require further study in order to define optimally discriminative cutoffs, but might facilitate better understanding of the condition by redirecting focus from the symptoms to the mechanisms by which they do or do not resolve.

In each symptom category of our survey, one participant reported symptoms lasting more than a year (a different participant in each category). This symptom duration was a clear outlier for each category. The participants reporting this duration for motor and sensory symptoms reported ‘a little’ and ‘slight’ functional limitation due to the symptoms respectively, while the participant reporting dissociation for more than a year endorsed ‘a lot’ for the symptom’s impact on their life. Given this, it is ambiguous whether the participants reporting long duration sensory and motor symptoms would have received a diagnosis of FND had they come to medical attention; however, it is likely that the participant with long duration dissociative symptoms would have met diagnostic criteria for depersonalization/derealization disorder.^12^ Under the schema we have suggested above, the participants with long duration functional neurological symptoms could more comfortably be categorized as having mild or subclinical FND. This would acknowledge both the clear difference between these symptoms and those experienced in normal life, and the substantial difference in symptom severity from people with more severe FND.

## Limitations

Our sample was deliberately chosen to include people we expected would be able to recognize the features of functional symptoms. Of necessity, however, this meant our sample might not have been representative of the wider population. The social and economic situations of healthcare professionals attending an international conference are likely to be favorable compared with those of many people, and it could reasonably be expected that the proportion of them affected by neurodevelopmental and mental health conditions known to be associated with FND might differ from that of the general population. Likewise, it is possible that some of the participants might have had their interest in FND stimulated by their experience of relevant symptoms. Conversely, perhaps some individuals experienced symptoms that were elicited by their exposure to people with FND. We cannot rule out possible biases introduced by these factors, however unlikely they might seem.

We included dissociation as a symptom of interest in our survey with the intention of capturing subclinical experiences within the spectrum of dissociative seizures, but it should be noted that the link between these experiences and dissociative seizures is less clear than that for the sensory and motor symptoms we asked participants to report. The sensorimotor symptoms that participants in our study reported share the key qualities of the symptoms which people with FND experience. By contrast, dissociation occurs commonly in people who have dissociative seizures, and more so in the peri-ictal period, but also occurs in other conditions, including as an isolated pathological process.^17,18^

## Summary

We have shown that functional symptoms occur commonly in people who do not have FND. Nosological formulations of FND need to be updated to acknowledge this, to better reflect the most likely pathological distinctions between benign functional symptoms and FND. Researchers should distinguish between research into the mechanisms of functional symptoms in general, and research into the mechanisms by which the brain remedies these symptoms, which may be more pertinent in the aetiology of FND.

## Funding

This work was supported by the Canadian Institute for Advanced Research (CIFAR). JBM was supported by a National Health and Medical Research Council (Australia) Investigator Grant (2010141).

## Competing interests

M.J.E. does medical expert reporting in personal injury and clinical negligence cases. M.J.E. has shares in Brain & Mind, which provides neuropsychiatric and neurological rehabilitation in the independent medical sector. M.J.E. has received financial support for lectures from the International Parkinson’s and Movement Disorders Society and the FND Society (FNDS). M.J.E. receives royalties from Oxford University Press for his book The Oxford Specialist Handbook of Parkinson’s Disease and Other Movement Disorder. M.J.E has received honoraria for medical advice to Teva Pharmaceuticals and educational events. M.J.E. receives grant funding from the National Institute for Health and Care Research (NIHR). M.J.E. is an associate editor of the European Journal of Neurology. M.J.E is a board member of the FNDS. M.J.E. is on the medical advisory boards of the charities FND Hope UK and the British Association of Performing Arts Medicine.

D.D.G.P and J.B.M have no competing interests to disclose.

## Data availability

The FNEQ is available as supplementary material at *CNS Spectrums* online.

