## Supplementary material for "Functional neurological symptoms occur commonly in healthy adults: implications for the pathophysiology of FND"

### Functional Neurological Experiences Questionnaire

1. Have you ever had trouble moving a body part in the way you wanted to (including swallowing) which got worse the harder you tried, or seemed to work better when you were not concentrating on it?
   *This includes both trouble getting a body part to move, and a body part moving in ways you didn't want or intend.*

   ☐ Yes ☐ No

   *If Yes, please answer 1a, 1b, and 1c below, otherwise continue to question 2.*

1a. How many times has this happened in your life?

- *Count each time a new symptom (or symptoms) began as a different instance.*
- *If a symptom was coming and going over short periods, count it as one instance.*
- *If a symptom resolved completely for a substantial amount of time then came back, count it as a different instance.*

| Number of times | Select one |
|---|---|
| Only once | ○ |
| 2 or 3 times | ○ |
| 4 – 6 times | ○ |
| 7 – 10 times | ○ |
| More than 10 times | ○ |

1b. How long did the longest of these symptoms last for?
*Count a symptom that was coming and going over short timeframes as one instance of the symptom.*

| Number of times | Select one |
|---|---|
| Less than a minute | ◯ |
| A few minutes | ◯ |
| More than a few minutes, but less than an hour | ◯ |
| More than an hour, but less than a day | ◯ |
| One day to one week | ◯ |
| One week to one month | ◯ |
| One month to three months | ◯ |
| Three months to a year | ◯ |
| More than a year | ◯ |

1c. At their worst, how much did these symptoms impact on your work, education, or social life?

| Number of times | Select one |
|---|---|
| Not at all | ◯ |
| Slightly *(eg, felt uncomfortable at work or school, or while socialising)* | ◯ |
| A little *(made work, education, or socialising difficult)* | ◯ |
| Quite a lot *(required up to a few days off work or school, or limited ability to socialise in certain situations)* | ◯ |
| A lot *(required more than a few days off work or school, or significantly limited ability to socialise)* | ◯ |

2. Have you ever had "phantom" sensations that did not occur related to another medical condition?
*This includes both numbness and feelings of things that aren't there, eg pins and needles, vibrations, or wetness.*

☐ Yes ☐ No

*If Yes, please answer 2a, 2b, and 2c below, otherwise continue to question 3.*

2a. How many times has this happened in your life?

- *Count each time a new symptom (or symptoms) began as a different instance.*
- *If a symptom was coming and going over short periods, count it as one instance.*
- *If a symptom resolved completely for a substantial amount of time then came back, count it as a different instance.*

| Number of times | Select one |
|---|---|
| Only once | ○ |
| 2 or 3 times | ○ |
| 4 – 6 times | ○ |
| 7 – 10 times | ○ |
| More than 10 times | ○ |

2b. How long did the longest of these symptoms last for?
*Count a symptom that was coming and going over short timeframes as one instance of the symptom.*

| Number of times | Select one |
|---|---|
| Less than a minute | ○ |
| A few minutes | ○ |
| More than a few minutes, but less than an hour | ○ |
| More than an hour, but less than a day | ○ |
| One day to one week | ○ |
| One week to one month | ○ |
| One month to three months | ○ |
| Three months to a year | ○ |
| More than a year | ○ |

2c. At their worst, how much did these symptoms impact on your work, education, or social life?

| Number of times | Select one |
|---|---|
| Not at all | ◯ |
| Slightly *(eg, felt uncomfortable at work or school, or while socialising)* | ◯ |
| A little *(made work, education, or socialising difficult)* | ◯ |
| Quite a lot *(required up to a few days off work or school, or limited ability to socialise in certain situations)* | ◯ |
| A lot *(required more than a few days off work or school, or significantly limited ability to socialise)* | ◯ |

3. Have you ever had dissociation (derealization or depersonalisation)?
*This can include feeling like being in a dream while you are awake, like you are not quite in your own body, like you are not quite in the world, or that it is as though there is a veil between you and the world.*

☐ Yes ☐ No

*If Yes, please answer 3a, 3b, and 3c below, otherwise continue to question 3.*

*3a.* How many times has this happened in your life?

- *Count each time a new symptom (or symptoms) began as a different instance.*
- *If a symptom was coming and going over short periods, count it as one instance.*
- *If a symptom resolved completely for a substantial amount of time then came back, count it as a different instance.*

| Number of times | Select one |
|---|---|
| Only once | ◯ |
| 2 or 3 times | ◯ |
| 4 – 6 times | ◯ |
| 7 – 10 times | ◯ |
| More than 10 times | ◯ |

3b. How long did the longest of these symptoms last for?

*Count a symptom that was coming and going over short timeframes as one instance of the symptom.*

| Number of times | Select one |
|---|---|
| Less than a minute | ○ |
| A few minutes | ○ |
| More than a few minutes, but less than an hour | ○ |
| More than an hour, but less than a day | ○ |
| One day to one week | ○ |
| One week to one month | ○ |
| One month to three months | ○ |
| Three months to a year | ○ |
| More than a year | ○ |

3c. At their worst, how much did these symptoms impact on your work, education, or social life?

| Number of times | Select one |
|---|---|
| Not at all | ○ |
| Slightly *(eg, felt uncomfortable at work or school, or while socialising)* | ○ |
| A little *(made work, education, or socialising difficult)* | ○ |
| Quite a lot *(required up to a few days off work or school, or limited ability to socialise in certain situations)* | ○ |
| A lot *(required more than a few days off work or school, or significantly limited ability to socialise)* | ○ |
